# Alcohol consumption and heavy episodic drinking within different types of drinking occasion in Great Britain: An event-level latent class analysis

**DOI:** 10.1101/2023.08.31.23294881

**Authors:** John Holmes, Alessandro Sasso, Mónica Hernández Alava, Rita Borges Neves, Abigail K Stevely, Alan Warde, Petra S Meier

**Affiliations:** School of Health and Related Research, University of Sheffield, Sheffield, UK; European Commission, Joint Research Center (JRC), Ispra, Italy; Management School, University of Sheffield, Sheffield, UK; School of Social Sciences, University of Manchester, Manchester UK; MRC/CSO Social and Public Health Sciences Unit, University of Glasgow, Glasgow, UK

## Abstract

**Aims:** To update a previous typology of British alcohol drinking occasions using a more recent and expanded dataset and revised modelling procedure. To estimate the average consumption level, prevalence of heavy drinking, and distribution of total alcohol consumption and heavy drinking within and across occasion types.

**Design:** Cross-sectional latent class analysis of event-level diary data.

**Setting:** Great Britain, 2019.

**Cases:** 43,089 drinking occasions reported by 17,821 adult drinkers.

**Measurements:** The latent class indicators are characteristics of off-trade only (e.g. home), on-trade only (e.g. bar) and mixed trade (e.g. home and bar) drinking occasions. These describe companions, venue(s) and location, purpose, motivation, accompanying activities, timings, weekday, consumption volume in units (1 UK unit = 8g ethanol) and predominant beverage consumed.

**Findings:** Three latent class models identified four off-trade only occasion types (i.e. latent classes), eight on-trade only occasion types and three mixed-trade occasion types. Mean consumption per occasion varied between 4.4 units in *Family meals* to 17.7 units in *Big nights out with pre-loading*. Mean consumption exceeded ten units in all mixed-trade occasion types and in *Off-trade get togethers, Big nights out* and *Male friends at the pub*. Three off-trade occasion types accounted for 50.8% of all alcohol consumed and 51.8% of heavy drinking occasions: *Quiet drink at home alone, Evening at home with partner* and *Off-trade get togethers*. For thirteen out of fifteen occasion types, more than 25% of occasions involved heavy drinking. Conversely, 41.7% of *Big nights out* and 16.4% of *Big nights out with preloading* were not heavy drinking occasions.

**Conclusions:** Alcohol consumption varies substantially across and within fifteen types of drinking occasion in Great Britain. Heavy drinking is common in most occasion types. However, moderate drinking is also common in types often characterised as heavy drinking practices. Mixed-trade drinking occasions are particularly likely to involve heavy drinking.

## INTRODUCTION

Epidemiological research increasingly examines alcohol consumption at the occasion- or event-level [1]. This work draws on diverse disciplinary and methodological traditions to address topics including the levels of alcohol consumption associated with different types of drinking occasions, the population groups that participate in those occasions, and the occasion characteristics that are associated with heavy drinking and harmful outcomes [1-7]. The findings offer a distinct perspective on alcohol consumption to inform public debate, engage stakeholders and guide prevention strategies [8]. In particular, they shift attention away from the characteristics of individual drinkers and instead focus on the wider social, commercial and environmental forces that shape alcohol consumption.

However, few event-level studies use nationally representative data [1]. This reflects the resource-intensive nature of event-level designs, such as diary studies, ecological momentary assessment and street intercept surveys, which often impose considerable burdens on researchers or their subjects [9-12]. Event-level studies often rely instead on small or unrepresentative samples that are temporally- or spatially-concentrated. There are, however, some exceptions. Paradis et al. used occasion-level data from a nationally representative Canadian telephone survey to show that reduced heavy drinking in parenthood may arise from fewer opportunities to drink in high consumption contexts, rather than lower consumption per occasion [13, 14]. Finnish researchers used a latent class analysis (LCA) of survey data on participants’ most recent drinking occasions to develop an occasion typology [5, 6]. They found that 40% of all reported alcohol consumption was within the 54% of occasions characterised as *At home with family* or *Home alone*, while *Big party nights* accounted for 11% of all drinking occasions but 30% of heavy drinking occasions. We conducted a similar typological analysis for Great Britain using one-week diary data and identified eight predominant occasion types, including *Drinking at home alone* (13.4% of occasions), *Drinking at home with family* (12.8% of occasions) and *Sociable get-togethers at someone’s house* (14.4% of occasions) [4]. Stevely et al. used the same diary data for 2018 to show that accounting for the common ways in which characteristics of occasions combine explains more variance in alcohol consumption than treating each characteristic as an independent variable [15].

Given the sparse literature in this area, further development and updating of such studies is important to provide basic epidemiological surveillance of event-level alcohol consumption. This paper therefore updates and extends our previous typology of British drinking occasions in 2009 to 2011 to address four key limitations [4]. First, it uses data from 2019 since there have been marked shifts in British drinking since 2011 [16-18]. Second, our previous analysis estimated a single latent class model that included all off-trade (e.g. home), on-trade (e.g. pub, restaurant or nightclub) and mixed-trade (e.g. home and pub) occasions. This limited the model’s ability to differentiate occasion types within each trade sector and meant we only identified one mixed-trade and two on-trade occasion types. As additional on-trade occasion types are readily identifiable within public discourse (e.g. big nights out, Sunday pub lunches, dinner dates), we now estimate separate models for each trade sector to better capture the diversity of drinking within these trade sectors. Third, our 2019 dataset includes information on additional occasion characteristics (e.g. food consumption and accompanying activities such as watching TV) that permit better differentiation of occasions within the latent class model. Fourth, we undertake secondary analyses of the LCA results to deepen understanding of the occasion types associated with greater levels of alcohol-related harm.

Specifically, this paper aims to estimate a new typology of the predominant drinking occasions observed in Great Britain during 2019 using an expanded dataset and improved modelling procedure. It then aims to estimate the average consumption level and prevalence of heavy drinking associated with each occasion type and the proportion of total reported alcohol consumption and heavy drinking occasions within each occasion type.

## METHODS

### Data

The data come from Alcovision, a continuous, cross-sectional, online study that surveys approximately 30,000 adults (18+) resident in Great Britain each year. The data are collected for market research purposes by Kantar, who draw quota samples each week based on age, gender, socioeconomic status and geographic region from their online managed access panels. Invitations to participate are timed to ensure that fieldwork includes every day of the year. Alcovision oversamples residents of Scotland and 18-34 year-olds to allow detailed analyses of these smaller populations. Kantar then construct sampling weights based on age-gender groups, social class and geographic region using UK census data. We have updated these weights using a bespoke raking technique [19].

Alcovision’s main component is a one-week, retrospective drinking diary in which participants report the characteristics of up to two on-trade and two-off-trade drinking occasions per day. This study uses a complete case analysis of the 2019 Alcovision data, which includes 43,089 drinking occasions reported by 17,821 individuals who consumed alcohol in the diary week.

## Measures

### Occasion trade sector

Kantar define an occasion as a significant time-period (e.g. lunchtime, late evening) within either the on-trade or off-trade. However, as familiar occasions such as ‘pre-drinking before a night out’ span both trade sectors, we use an alternative definition to better characterize an occasion. Using the reported start-time and duration of each occasion in Alcovision, we specify an occasion as a period of alcohol consumption with no more than two hours between drinks [5]. This allows us to separate occasions into three categories: off-trade only, on-trade only and mixed-trade (i.e. occasions involving both trade sectors).

### Occasion characteristics

Participants record detailed information about each drinking occasion in a series of categorical variables that are summarised below and described fully in the appendix Table A1. We have previously argued that these variables capture key concepts that are important to a social practice perspective on alcohol consumption [4].

Two measures capture the people present: the *sex composition of the participants* (e.g. male alone, female pair) and who any *companions* were (e.g. family, friends). Four measures capture on-trade locations: the *venue* (e.g. own home, modern bar), the *venue location* (e.g. village or rural, city centre), the *reasons for choosing the venue* (e.g. friendly atmosphere, cheap) and a dichotomous variable for visiting *multiple on-trade venues*. Two measures capture the nature of the occasion: the *purpose* (e.g. quiet night in, going clubbing) and the *motivation* for the occasion (e.g. to wind down, to have a laugh). Two measures capture activities accompanying drinking: *general activities* (e.g. watching TV, doing a pub quiz) and *food eaten* (no food, a snack, a meal). Four measures capture the occasion temporalities: the *duration* (e.g. <1 hour, 4-7 hours), the *start-time* (e.g. lunchtime, night-time), the *day of the week* (Monday to 17:00 on Friday, 17:00 on Friday to 23:59 on Saturday, Sunday) and the *sequencing* of off-trade and on-trade drinking in mixed-trade occasions (e.g. pre-loading [off then on], post-loading [on then off]). Two measures capture alcohol consumption: *volume consumed* based on units (1 UK unit = 8g ethanol; e.g. 0.0-2.0 units, 5.0-12.0 units) and the *predominant beverage type*, which the respondent reports consuming most servings of (e.g. beer, wine).

### Summary consumption measures

The summary measures describe the mean and standard deviation of units consumed in each occasion type, and the percentage of all units, off-trade units, on-trade units and units of each beverage type observed in the dataset that are consumed in each occasion type. We also calculate the percentage of all occasions within each type that involve heavy drinking and the percentage of all observed heavy drinking occasions that are within each type. Heavy drinking occasions are identified using the standard UK definition of >6 units (48g) for women and >8 units (64g) for men.

## Analysis

The analysis has four stages. First, a description of the dataset using summary statistics. Second, estimation of the LCA model. Third, assignment of observed occasions to latent classes (i.e. occasion types). Fourth, calculation of the summary consumption measures for the observed occasions in each class. The first and fourth stages are descriptive analyses, so we focus on the second and third stages below.

We estimated separate LCA models for off-trade only, on-trade only and mixed-trade occasions. LCA assumes that the cases in a dataset can be separated into different classes (i.e. drinking occasion types) that are ‘latent’ as they cannot be observed directly. It uses variables describing each case (i.e. drinking occasion characteristics) to infer which cases may belong to each class together with a model that estimates the probability each case belongs to each class. The model uses appropriate parametric specifications for each occasion characteristic variable; specifically probit models for binary variables, ordered probit models for ordinal variables, and multinomial logit models for nominal variables. The probability of class membership is modelled using a multinomial logit model. The analyst iteratively determines the number of classes the LCA model should identify by assessing the impact of additional classes on model fit and interpretability. We used the Bayesian Information Criterion (BIC), Sample-Size Adjusted BIC, Akaike’s Information Criterion, and model entropy as standard goodness of fit measures, and also used the Vuong-Lo-Mendell-Rubin likelihood ratio test (LRT), the Lo-Mendell-Rubin adjusted LRT and the parametric bootstrapped LRT to test whether each additional class improved model fit. These tests require careful interpretation as they are sensitive to the sample size and number of model parameters, both of which are large in Alcovision [20]. This meant we particularly relied on qualitative assessments of interpretability using the research team’s topic expertise and in consultation with our project’s expert advisory group. After estimating the final occasion typology, we deterministically assigned each occasion to a latent class based on predicted posterior probabilities.

All models used weighted data and also included clustered standard errors to account for the nesting of occasions within individuals. The LCA models were estimated via maximum likelihood using Mplus v8.3.0.1. The analyses were not pre-registered and should be considered exploratory.

## RESULTS

### Descriptive analysis

Table 1 provides summary statistics for selected variables (see Appendix 2, Table A2.1 for all variables). Approximately two-thirds (68.9%) of drinking occasions in Great Britain in 2019 were off-trade only, 20.8% were on-trade only and 10.3% were mixed-trade. The mean number of units consumed was lowest in off-trade only occasions at 7.2 units, compared to 8.1 units in on-trade only occasions and 14.9 units in mixed-trade occasions. Off-trade only occasions were also less likely to involve heavy drinking than on-trade only or mixed-trade occasions. However, off-trade only occasions still accounted for 58.2% of all alcohol consumed and 59.7% of heavy drinking occasions. In contrast, mixed-trade occasions accounted for 20.4% of all alcohol consumption, 76.6% involved heavy drinking and these accounted for 19.3% of all heavy drinking occasions.

**Table 1:** Summary statistics for occasion characteristics and alcohol consumption for drinking occasions in Great Britain, 2019.

| Characteristic<sup>1</sup> | Occasions |  | Units consumed |  |  | % of total consumption<sup>2</sup> |  |  | % that are HDO<sup>3</sup> | % of all HDO |
| --- | --- | --- | --- | --- | --- | --- | --- | --- | --- | --- |
|  | N | % | Mean | SD | Median | Total | Off | On |  |  |
| All | 43,089 | 100.0 | 8.2 | 8.4 | 5.3 | 100.0 | 100.0 | 100.0 | 41.1 | 100.0 |
| **Trade sector** |  |  |  |  |  |  |  |  |  |  |
| Off-trade | 29,678 | 68.9 | 7.2 | 7.6 | 4.2 | 58.2 | 84.1 | - | 35.6 | 59.7 |
| On-trade | 8,956 | 20.8 | 8.1 | 8.0 | 5.4 | 21.4 | - | 69.5 | 41.6 | 21.0 |
| Mixed | 4,455 | 10.3 | 14.9 | 10.8 | 12.0 | 20.4 | 15.9 | 30.5 | 76.6 | 19.3 |
| **Sex composition** |  |  |  |  |  |  |  |  |  |  |
| Mixed sex group | 16,630 | 37.0 | 10.6 | 10.1 | 7.0 | 52.1 | 47.2 | 63.0 | 49.2 | 47.9 |
| Mixed sex pair | 14,064 | 31.3 | 6.7 | 6.8 | 4.1 | 22.8 | 26.2 | 15.2 | 31.9 | 26.2 |
| Male alone | 5,187 | 11.6 | 7.8 | 7.8 | 9.1 | 10.6 | 11.8 | 7.9 | 34.3 | 10.4 |
| **Companions** |  |  |  |  |  |  |  |  |  |  |
| Partner | 19,518 | 43.5 | 7.8 | 8.0 | 5.1 | 39.2 | 42.9 | 30.9 | 37.1 | 42.3 |
| Friends | 12,069 | 26.9 | 11.7 | 10.2 | 8.4 | 42.7 | 34.8 | 60.3 | 55.4 | 39.1 |
| Family | 9,024 | 20.1 | 9.2 | 9.6 | 5.6 | 23.9 | 24.1 | 23.4 | 41.9 | 22.1 |
| **Venue** |  |  |  |  |  |  |  |  |  |  |
| Own home (off-trade) | 29,513 | 65.7 | 7.7 | 8.1 | 4.7 | 59.0 | 76.3 | 20.4 | 35.7 | 61.7 |
| Other's home (off-trade) | 4,506 | 10.0 | 11.8 | 10.7 | 8.0 | 16.4 | 20.4 | 7.4 | 54.1 | 14.3 |
| Traditional pub (on-trade) | 5,074 | 11.3 | 11.9 | 10.0 | 9.0 | 16.9 | 5.5 | 42.3 | 57.5 | 17.1 |
| Modern bar (on-trade) | 2,566 | 5.7 | 13.1 | 11.0 | 9.4 | 10.6 | 3.5 | 26.4 | 60.1 | 9.0 |
| Multiple venues (on-trade) | 2,182 | 4.9 | 17.5 | 11.8 | 6.1 | 12.5 | 3.4 | 32.9 | 78.6 | 10.0 |
| **Purpose of occasion** |  |  |  |  |  |  |  |  |  |  |
| Quiet drink (off-trade) | 11,146 | 24.8 | 7.9 | 7.8 | 5.3 | 23.0 | 30.7 | 5.9 | 37.8 | 24.7 |
| Regular drink (off-trade) | 11,032 | 24.6 | 7.7 | 8.0 | 4.7 | 20.6 | 27.6 | 5.0 | 35.4 | 22.8 |
| Other (on-trade) | 6,663 | 14.8 | 10.4 | 9.8 | 6.9 | 20.0 | 7.8 | 47.3 | 55.6 | 47.3 |
| Sociable (on-trade) | 2,870 | 6.4 | 11.1 | 9.4 | 8.3 | 8.8 | 3.0 | 21.8 | 49.0 | 21.8 |
| **Food eaten** |  |  |  |  |  |  |  |  |  |  |
| None | 17,806 | 39.7 | 7.3 | 7.3 | 4.5 | 34.4 | 29.6 | 45.2 | 33.8 | 35.2 |
| Snack | 6,880 | 15.3 | 8.7 | 8.8 | 5.5 | 16.9 | 18.3 | 13.8 | 41.1 | 16.6 |
| Meal | 20,207 | 45.0 | 8.8 | 9.1 | 5.5 | 48.7 | 52.1 | 41.0 | 40.8 | 48.3 |
| **Duration** |  |  |  |  |  |  |  |  |  |  |
| Less than 1 hour | 11,641 | 25.9 | 3.4 | 3.9 | 2.3 | 9.7 | 11.6 | 5.5 | 8.2 | 5.6 |
| 1-4 hours | 26,888 | 59.9 | 8.3 | 7.6 | 5.8 | 59.6 | 59.9 | 59.0 | 41.7 | 65.7 |
| 4-7 hours | 5,418 | 12.1 | 15.8 | 10.6 | 13.7 | 24.9 | 23.1 | 29.0 | 77.0 | 24.4 |
| **Start-time** |  |  |  |  |  |  |  |  |  |  |
| Evening | 26,922 | 60.0 | 7.9 | 7.9 | 5.3 | 56.8 | 60.2 | 49.1 | 38.0 | 59.9 |
| Afternoon | 9,360 | 20.8 | 9.6 | 9.6 | 5.8 | 24.6 | 23.3 | 27.5 | 43.6 | 23.9 |
| Lunchtime | 3,718 | 8.3 | 8.1 | 9.1 | 4.5 | 7.9 | 6.5 | 11.0 | 34.2 | 7.4 |
| **Day of week** |  |  |  |  |  |  |  |  |  |  |
| Mon – Fri (17:00) | 17,175 | 38.3 | 7.4 | 7.9 | 4.4 | 34.8 | 34.6 | 35.3 | 33.2 | 33.3 |
| Fri (17:00) – Sat | 20,744 | 46.2 | 8.9 | 8.7 | 5.7 | 50.5 | 50.1 | 51.2 | 42.5 | 51.6 |
| Sunday | 6,974 | 15.5 | 8.0 | 8.4 | 5.0 | 14.7 | 15.2 | 13.4 | 37.0 | 15.1 |
| **Units of alcohol consumed** |  |  |  |  |  |  |  |  |  |  |
| 0.0 – 2.0 | 7,554 | 16.8 | 1.4 | 0.4 | 1.4 | 2.5 | 2.9 | 1.6 | 0.0 | 0.0 |
| 2.0 – 3.5 | 8,249 | 18.4 | 2.5 | 0.4 | 2.7 | 5.1 | 5.4 | 4.5 | 0.0 | 0.0 |
| 3.5 – 5.0 | 6,204 | 13.8 | 4.1 | 0.4 | 4.0 | 6.1 | 6.3 | 5.5 | 0.0 | 0.0 |
| 5.0 – 12.0 | 13,745 | 30.6 | 8.0 | 2.0 | 7.9 | 28.1 | 28.8 | 26.5 | 57.9 | 46.5 |
| 12.0 – 20.0 | 5,069 | 11.3 | 15.6 | 2.3 | 15.3 | 22.0 | 21.3 | 23.4 | 100.0 | 29.7 |
| > 20.0 | 4,071 | 9.1 | 29.7 | 7.1 | 28.3 | 36.3 | 35.2 | 38.5 | 100.0 | 23.8 |
| **Predominant beverage** |  |  |  |  |  |  |  |  |  |  |
| Beer | 15,702 | 35.0 | 8.0 | 7.9 | 43.1 | 33.1 | 28.7 | 43.1 | 37.0 | 34.0 |
| Wine | 12,976 | 28.9 | 7.9 | 7.6 | 14.5 | 24.4 | 28.8 | 14.5 | 40.2 | 30.5 |
| Spirits | 10,584 | 23.6 | 9.2 | 10.0 | 28.2 | 29.6 | 30.3 | 28.2 | 39.9 | 24.7 |

<sup>1</sup>Selected variables and highest prevalence categories within each variable only. See Appendix Table A2.1 for full data. Categories labelled off-trade or on-trade relate to those trade sectors only. <sup>2</sup>Percentage of overall consumption in trade sector occurring in occasions with this characteristic. Occasions with off-trade characteristics account for a proportion of on-trade consumption (and vice versa) if they are mixed-trade occasions. <sup>3</sup>HDO: Heavy drinking occasions.

### Latent class analyses

We estimated models with between two and eight latent classes for each trade sector (see Appendix 1, Figures A1.1-A1.3 and Table A1.1 for model fit statistics). The off-trade only model showed decreasing improvements in most fit statistics (e.g. AIC, BIC, adjust-BIC) and limited interpretability beyond four classes. The on-trade only model showed continued improvements in model fit and interpretability up to eight classes, but did not converge when identifying further classes due to low frequencies in some of the observed variables. The mixed-trade models showed limited interpretability beyond three classes and no improvements in model fit beyond four or five classes using the Vuong-Lo-Mendell-Rubin LRT and Lo-Mendell-Rubin adjusted LRT. We therefore selected final models with four latent classes for off-trade only occasions, eight latent classes for on-trade only occasions, and three latent classes for mixed-trade occasions. The entropy value was above 0.8 for all classes. We reran the alcohol consumption analyses excluding the 9.5% of occasions with a class membership probability below 0.8 but this did not substantively affect our results, so we include all occasions in the results below.

Figure 1 reports selected characteristics for each class (see Appendix 2, Table A2.2 for full results). The descriptions in Figure 1 and below do not discuss alcohol consumption levels as the final section of the analysis addresses this and we encourage readers to focus first on the wider characteristics of each occasion type. For clarity, we refer to latent classes as occasion types henceforth.

**Figure 1:**
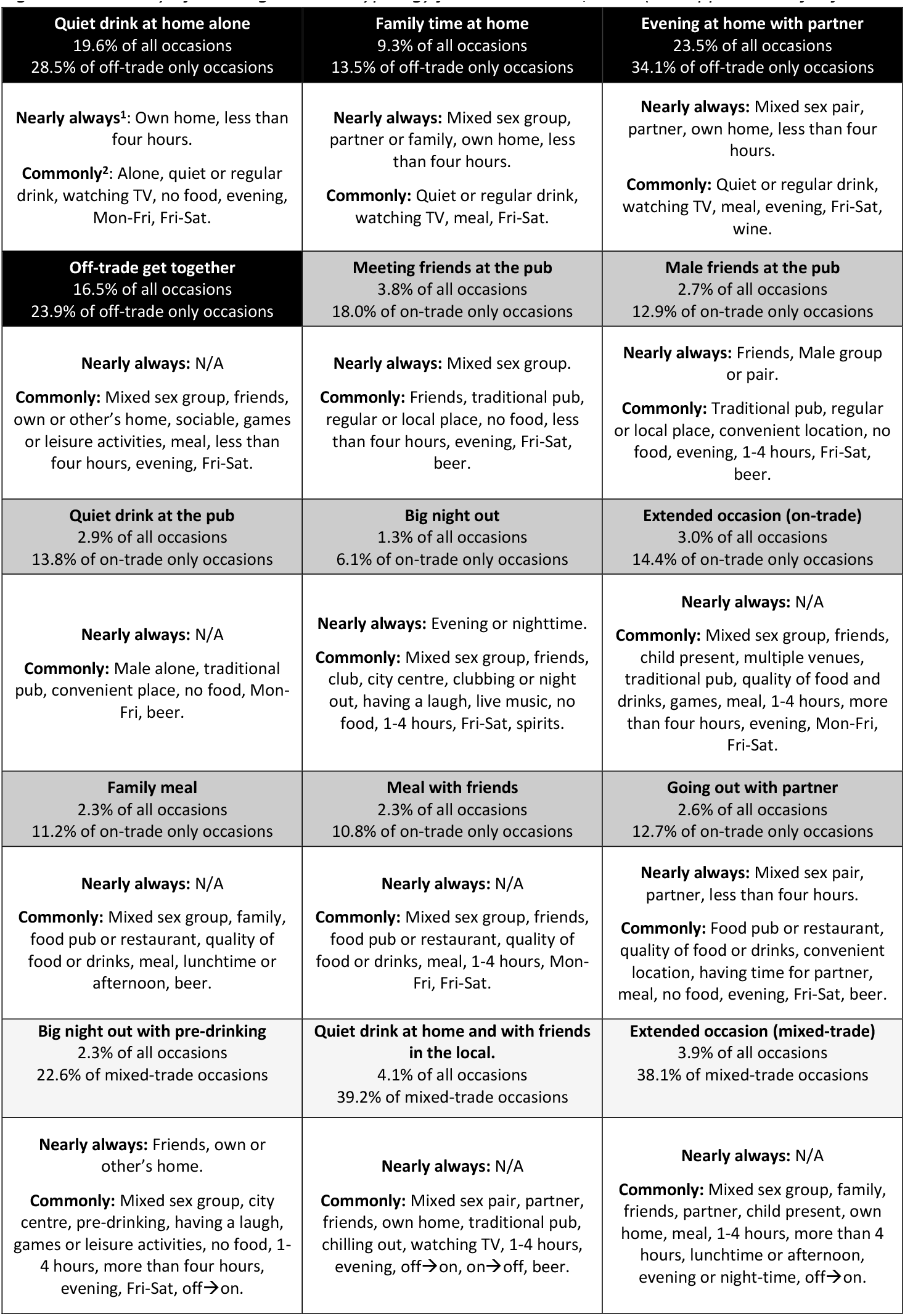
Summary of drinking occasion typology for Great Britain, 2019 (see Appendix A2 for full results).

Three of the four off-trade only occasion types involve drinking with other household members and jointly account for 76.1% of off-trade only occasions. We labelled these: *Quiet drink at home alone, Family time at home*, and *Evening at home with partner*. These are quiet, regular occasions accompanied by low-key activities, such as eating a meal or watching TV. They are typically short, in the evening and occur throughout the week, but particularly at weekends. The remaining off-trade only occasion type is *Off-trade get together*. It involves larger groups of friends and family, an emphasis on sociability, and playing games or other leisure activities. These occasions are often longer, more likely to start earlier in the day and more concentrated on weekends.

The on-trade only occasion types are more diverse and can be grouped into pub-drinking occasions, longer occasions, and meals or dates. The three pub-drinking occasions jointly account for 44.7% of on-trade only occasions. We labelled them: *Meeting friends at the pub, Male friends at the pub* and *Quiet drink at the pub*. They involve beer-drinking in traditional pubs or similarly convenient, local or regular venues and are unlikely to involve a meal or other accompanying activities. The *Quiet drink at the pub* type is lower-key than the other two occasion types, more often involves males alone, is shorter, and more likely to occur earlier in the day.

The two longer occasions often involve multiple venues. These jointly account for 20.5% of on-trade only occasions and we labelled them: *Big night out* and *Extended occasion (on-trade). Big nights out* typically involve groups of friends having an upbeat outing in lively bars or clubs with music or dancing in city centres on a weekend evening. Spirits are often the dominant beverage. *Extended occasions (on-trade)* share these characteristics but are more heterogeneous. They additionally involve edge of town retail or entertainment complexes, take place on weekdays, start earlier in the day, and often involve a meal and activities such as dancing, karaoke or barroom games. They are also more likely to involve family members and children.

The remaining three on-trade only occasion types are *Family meal, Meal with friends* and *Date with partner* and account for 34.8% of on-trade only occasions. *Family meals* typically involve family, partners and children in food-serving venues at lunchtime, afternoon or evening on all days, with wine more commonly the dominant beverage than in the occasion types above. *Meals with friends* share many of these characteristics, excepting friends being present, a greater emphasis on socialising and a lower likelihood of taking place on Sundays. *Going out with partner* occasions are also similar but are more likely to involve mixed sex pairs, spending time together and a wider range of venues, although food-serving venues and meals remain common.

The three mixed-trade occasion types are distinct from each other and often include significant diversity within each type. *Big nights out with pre-drinking* share the characteristics of their on-trade only equivalent, but start earlier in the day and often involve pre-drinking at someone’s home while getting ready and participating in games and other leisure or online activities. Bars and nightclubs in city centres remain common venues but these mixed-trade nights out are more likely to involve traditional pubs and less likely to involve dancing and live music, or eating a meal or snack. *Quiet* d*rinks at home and with friends in the local* are generally, but not exclusively, low-key occasions that involve socialising but also spending quality time with family or alone at home and in traditional pubs that are convenient, local or friendly. Finally, *Extended occasions (mixed-trade)*, like their on-trade counterpart, appear fluid and heterogenous, variously involve groups of friends, family and sometimes children, having an upbeat occasion across a range of venues and over longer time periods starting at any time of day.

### Analysis of alcohol consumption

Table 2 presents descriptive statistics on the alcohol consumption associated with each occasion type (see Appendix 2, Table A2.3 for units by beverage type). The mean units consumed by respondents varies across occasion types from 4.4 units in *Family meals* to 17.7 units in *Big nights out with pre-drinking*. It is markedly higher in *Off-trade get togethers* (10.4 units) compared to other off-trade only occasions (5.9-6.9 units). It is also higher in *Mixed-trade extended occasions* (15.9 units), *Big nights out* (11.1 units) and *Male friends at the pub* (10.2 units) occasions compared to other on-trade only occasions (4.4-7.4 units). Mean consumption is over 12 units in all mixed-trade occasion types.

**Table 2:** Levels and distribution of alcohol consumption and heavy drinking occasions across occasion types (latent classes).

| Trade sector | Occasion type | % of occasions |  | Units consumed in occasion |  |  |  |  |  |  |  |  | % of total consumption |  |  | Heavy drinking occasions (HDOs) |  |
| --- | --- | --- | --- | --- | --- | --- | --- | --- | --- | --- | --- | --- | --- | --- | --- | --- | --- |
|  |  | All | In trade sector | Total |  |  | Off-trade |  |  | On-trade |  |  | Total | Off-trade | On-trade | % of type<sup>1</sup> | % of all HDOs |
|  |  |  |  | Mean | SD | Median | Mean | SD | Median | Mean | SD | Median |  |  |  |  |  |
| Off-trade only | Quiet drink at home alone | 19.6 | 28.5 | 6.2 | (6.7) | 3.8 | 6.2 | (6.7) | 3.8 | - | - | - | 13.2 | 19.1 | - | 27.3 | 14.1 |
|  | Family time at home | 9.3 | 13.5 | 6.9 | (7.0) | 4.2 | 6.9 | (7.0) | 4.2 | - | - | - | 7.3 | 10.6 | - | 32.3 | 7.9 |
|  | Evening at home with partner | 23.5 | 34.1 | 5.9 | (5.8) | 4.0 | 5.9 | (5.8) | 4.0 | - | - | - | 14.0 | 20.3 | - | 27.4 | 16.9 |
|  | Off-trade get together | 16.5 | 23.9 | 10.4 | (10.0) | 6.8 | 10.4 | (10.0) | 6.8 | - | - | - | 23.6 | 34.2 | - | 48.1 | 20.8 |
| On-trade only | Meeting friends at the pub | 3.8 | 18.0 | 7.4 | (6.1) | 5.7 | - | - | - | 7.4 | (6.1) | 5.7 | 3.3 | - | 10.6 | 40.4 | 4.0 |
|  | Male friends at the pub | 2.7 | 12.9 | 10.2 | (6.7) | 9.1 | - | - | - | 10.2 | (6.7) | 9.1 | 2.9 | - | 9.3 | 55.7 | 3.9 |
|  | Quiet drink at the pub | 2.9 | 13.8 | 5.5 | (4.6) | 4.1 | - | - | - | 5.5 | (4.6) | 4.1 | 1.9 | - | 6.0 | 23.5 | 1.8 |
|  | Big night out | 1.3 | 6.1 | 11.1 | (9.1) | 8.5 | - | - | - | 11.1 | (9.1) | 8.5 | 2.3 | - | 7.5 | 58.3 | 1.9 |
|  | Extended occasion (on-trade) | 3.0 | 14.4 | 15.9 | (12.2) | 13 | - | - | - | 15.9 | (12.2) | 13 | 7.0 | - | 22.7 | 68.9 | 5.4 |
|  | Family meal | 2.3 | 11.2 | 4.4 | (4.3) | 2.8 | - | - | - | 4.4 | (4.3) | 2.8 | 1.2 | - | 3.9 | 17.0 | 1.0 |
|  | Meal with friends | 2.3 | 10.8 | 5.3 | (5.3) | 3.8 | - | - | - | 5.3 | (5.3) | 3.8 | 1.4 | - | 4.6 | 24.1 | 1.4 |
|  | Going out with partner | 2.6 | 12.7 | 5.1 | (4.8) | 3.2 | - | - | - | 5.1 | (4.8) | 3.2 | 1.5 | - | 4.8 | 22.5 | 1.6 |
| Mixed-trade | Big night out with pre-drinking | 2.3 | 22.6 | 17.7 | (10.8) | 15.2 | 8.7 | (7.5) | 6.4 | 9.0 | (7.2) | 7.4 | 6.0 | 4.4 | 9.7 | 83.6 | 5.1 |
|  | Quiet drink at home and with friends... | 4.1 | 39.2 | 12.1 | (8.5) | 10.2 | 6.1 | (5.9) | 4.1 | 6.0 | (5.2) | 4.5 | 5.5 | 4.1 | 8.8 | 65.2 | 7.0 |
|  | Extended occasion (mixed trade) | 3.9 | 38.1 | 16.2 | (12.3) | 13 | 9.3 | (9.2) | 5.7 | 6.9 | (7.0) | 4.5 | 8.8 | 7.4 | 12.1 | 69.6 | 7.2 |

<sup>1</sup>Percentage of occasions within type that are heavy drinking occasions

Respondents’ alcohol consumption varies substantially within occasion types (see standard deviations in Table 2) and heavy drinking is common in most types. For example, in thirteen out of fifteen types, more than 25% of occasions involved the respondent drinking heavily. Conversely, respondents did not drink heavily on 41.7% of *Big night out* occasions and 16.4% of *Big night out with pre-drinking* occasions, which are stereotypically considered heavy drinking occasion types.

Half (50.8%) of all alcohol consumed and more than a third of heavy drinking occasions (51.8%) were in just three occasion types: *Quiet drink at home, Evening at home with partner* and *Off-trade get together* (Figure 2). The three mixed-trade types account for a further 20.4% of consumption and 19.3% of heavy drinking occasions, despite comprising only 10.3% of occasions. Notably, *Big nights out* account for just 2.3% of total consumption and 1.9% of heavy drinking occasions, while *Big nights out with pre-drinking* account for just 6.0% of total consumption and 5.1% of heavy drinking occasions.

**Figure 2:**
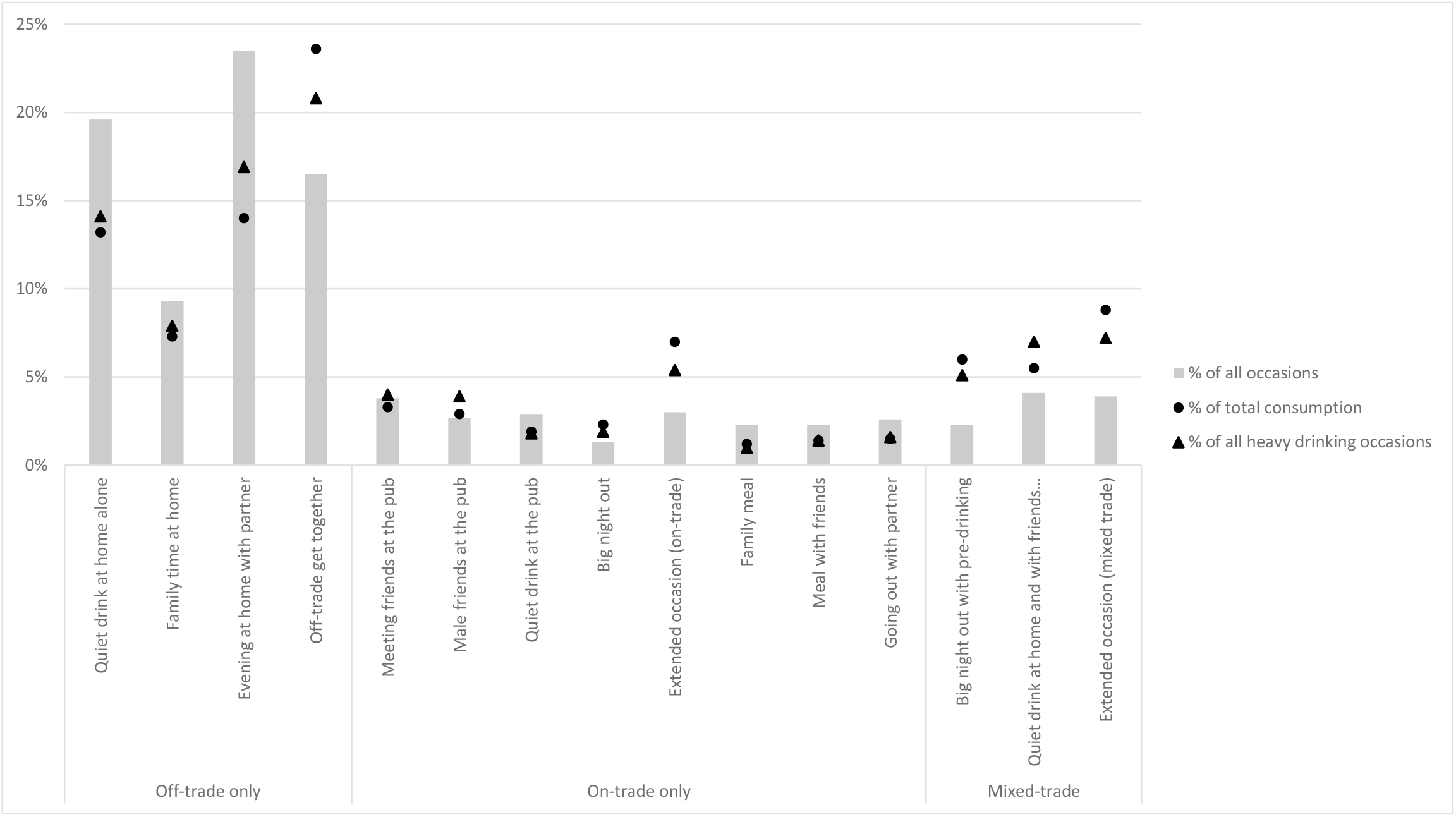
Distribution of drinking occasions, total alcohol consumption and heavy drinking occasions across occasion types estimated from latent class models.

Finally, Figure 3 demonstrates that substantial proportions of heavy drinking occasions are in occasion types with lower mean consumption. For example, 48.7% of heavy drinking occasions are in the eight occasion types where mean consumption was less than eight units and 38.9% of heavy drinking occasions are in the three off-trade only types with mean consumption below seven units.

**Figure 3:**
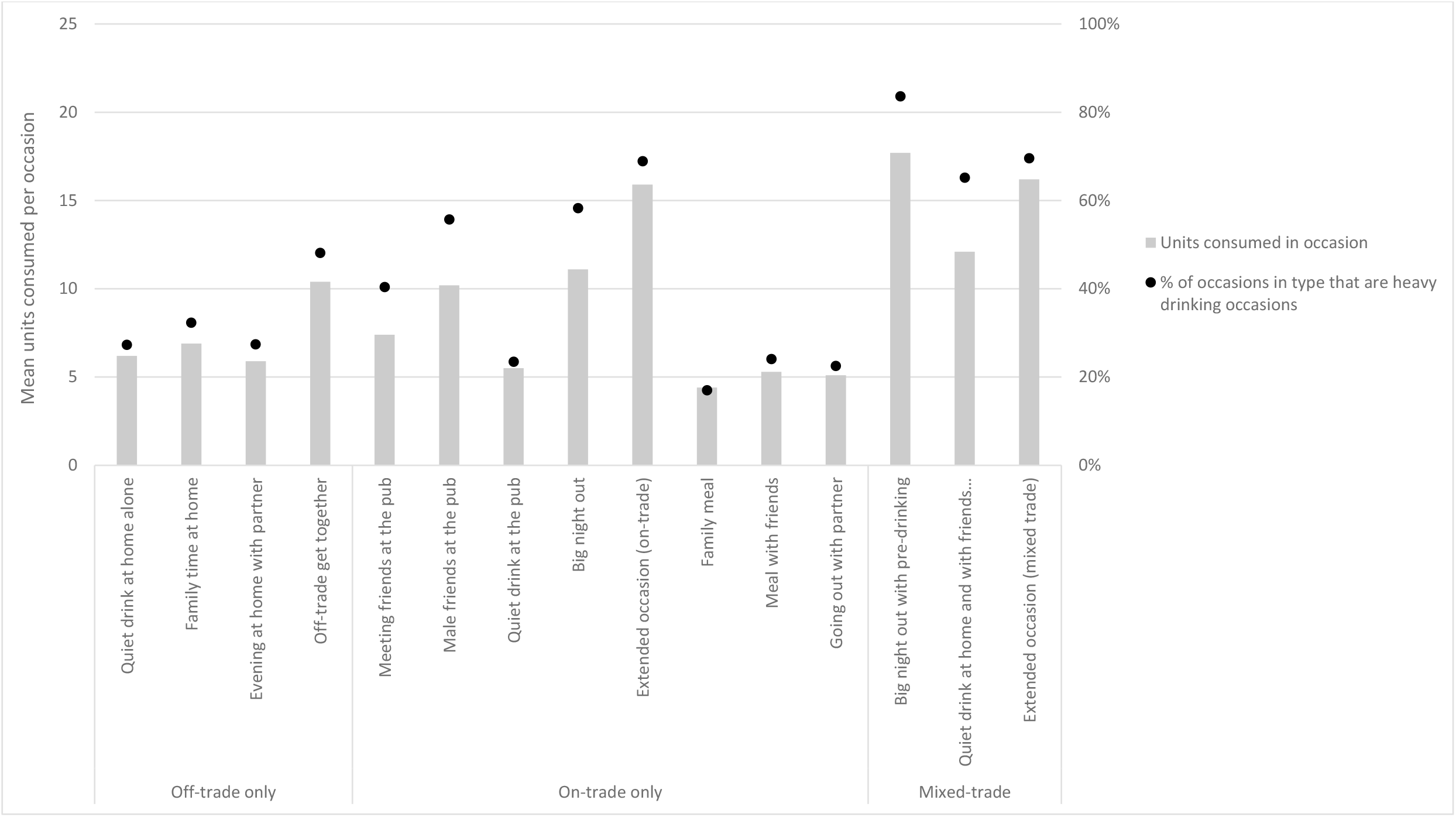
Mean units consumed by occasion type and proportion of occasions in type that are heavy drinking occasions estimated from latent class models.

Figure 4 and 5 present sex-specific analyses of consumption across occasions (see also Appendix 1, Tables A1.3 and A1.4). A greater proportion of women’s occasions, consumption and heavy drinking occasions are in the off-trade and particularly in *Evening at home with partner* or *Off-trade get together* occasions. A greater proportion of men’s occasions involve heavy drinking for all occasion types.

**Figure 4:**
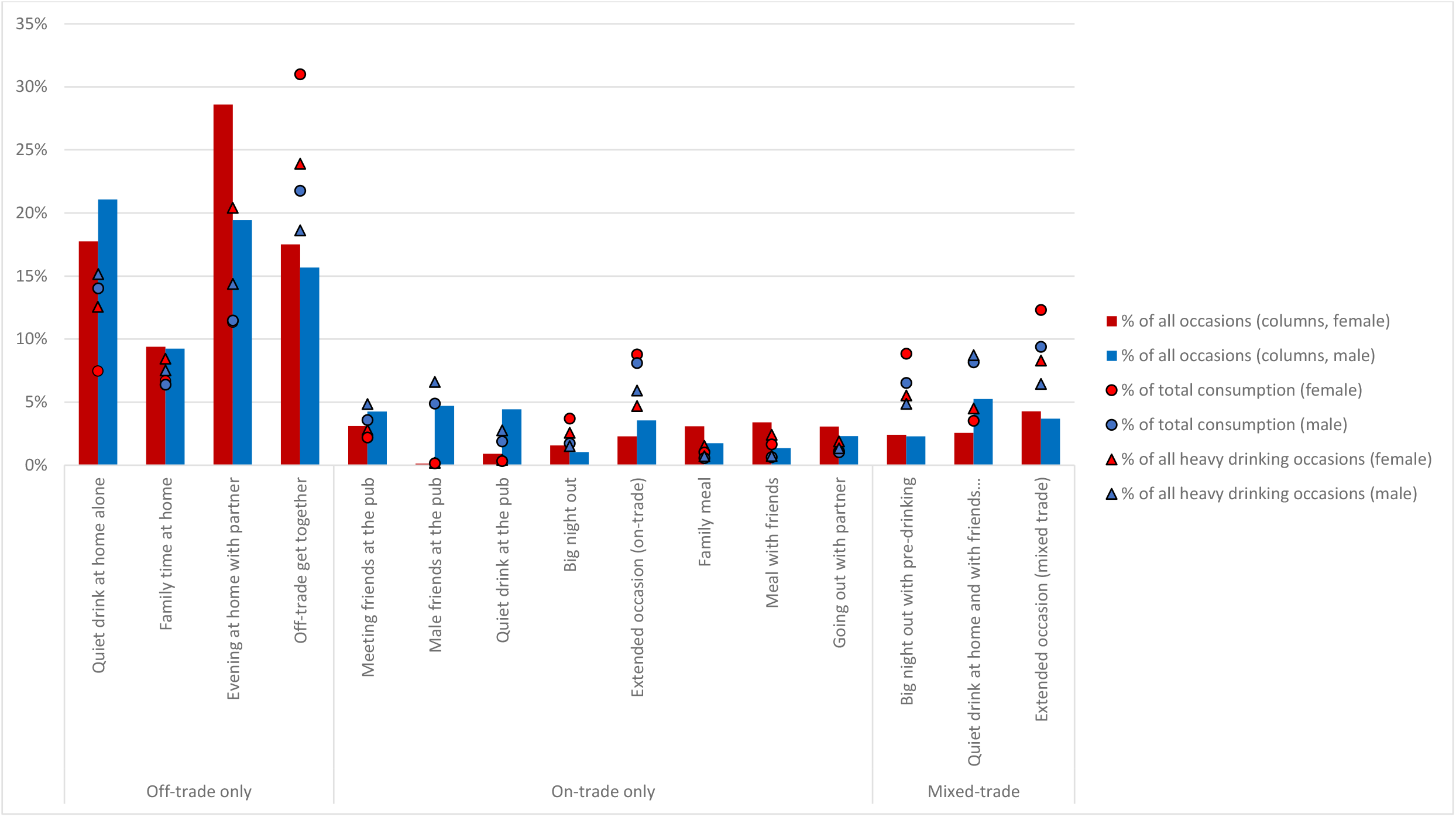
Distribution of drinking occasions, total alcohol consumption and heavy drinking occasions across occasion types by sex estimated from latent class models.

**Figure 5:**
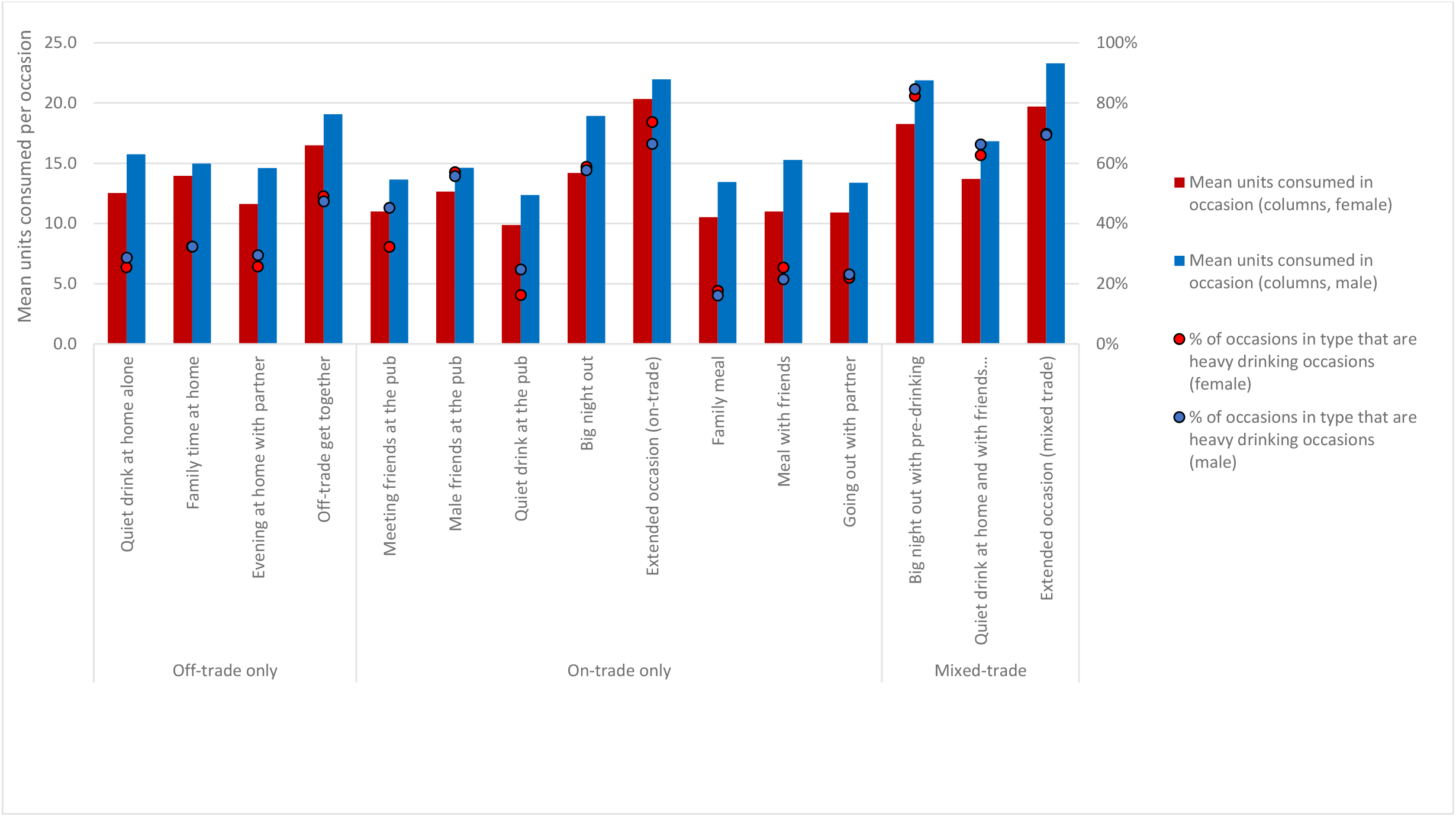
Mean units consumed by occasion type and proportion of occasions in type that are heavy drinking occasions by sex estimated from latent class models.

## Discussion

We used three latent class models to identify 15 types of drinking occasion in Great Britain. The four off-trade only occasion types were mostly low-key and only differed in minor ways from one another, except for the more lively *Off-trade get togethers*. The minor distinctions are nonetheless important as they reflect topics of public health relevance, such as drinking alone and drinking with children. The eight on-trade only types were diverse and include occasions of varying sociability in local pubs, lively days or nights out, family occasions and time spent with romantic partners. The three mixed-trade types are even more diverse and merit scrutiny because they involve high levels of alcohol consumption.

The findings suggest that alcohol consumption varies substantially across and within occasion types. Off-trade only occasion types account for a majority of alcohol consumption and heavy drinking occasions, but heavy drinking is common across almost all occasion types and is not concentrated within particular problematic types such as *Big nights out*. Indeed, a large proportion of heavy drinking takes place in occasion types such as *Quiet drink at home alone* and *Evening at home with partner*, which are generally commonly viewed as low-key or moderate. Conversely, people often drink moderately in occasion types usually assumed to involve heavy drinking, such as *Big nights out*. Few previous studies have examined the extent of moderate drinking within stereotypically heavy drinking occasion types, except when examining specific reasons for moderation, such as being a designated driver, a permanent abstainer or looking after intoxicated peers [21-23]. Qualitative research in particular emphasises narratives of excess within such occasions, particularly among young adults [24-27]. Our novel findings may reflect our focus on a wider age range than previous studies, or perhaps the tendency for policy debate to focus on visible public intoxication rather than unseen private intoxication.

Mixed-trade drinking occasions also appear important for understanding harmful alcohol consumption as at least 65% of occasions involved heavy drinking in all three mixed-trade occasion types. Event-level studies identify occasions involving drinking across the on- and off-trade trade sectors as commonplace, associated with higher consumption levels and therefore higher risk [4, 15, 28, 29]. However, beyond the specific practice of pre-drinking among young adults [30-32], little is known about the varying forms such occasions may take, who engages in them and how they contribute to some drinkers reaching high levels of alcohol consumption in a single day or across a week. Our findings suggest that some mixed-trade occasions are a distinct type of event (e.g. *Big nights out with pre-drinking)*, but others are more nebulous and may reflect individuals participating in several types of bounded occasion across a day (e.g. a family meal followed by meeting friends at the pub). The latter form of mixed-trade occasion may be particularly importance for prevention research if it facilitates heavy alcohol consumption while avoiding the informal social controls that limit drinking within a single occasion.

A key strength of our analysis is our large, detailed and nationally-representative dataset, which increases the robustness of the LCA models and permits greater disaggregation of occasion types in each trade sector. Our analysis also examines mixed-trade occasions, which capture commonplace movements of individuals between trade sectors that are not always incorporated into event-level studies. The analysis does however have important limitations. Alcovision is a market research study designed primarily to provide timely information to commercial clients. The sampling strategy and measures therefore have important limitations that we discuss elsewhere (e.g. the use of quota sampling and unvalidated measures that do not address key public health concerns) [4, 33]. These points may limit the representativeness of our typology to the extent early responders to invitations within market research panels differ from the general population.

The discussion above has three key implications for policy and practice. First, prevention strategies focused primarily on stereotypically heavy drinking practices may not impact the large proportion of heavy drinking that takes place within other practices. Broader strategies are required that recognise heavy drinking occurs routinely within most drinking practices and all trade sectors. Second, promoting moderate drinking within stereotypically heavy drinking practices may still reduce harm, as heavy drinking does not appear an intrinsic element of such practices for a large proportion of drinkers. Third, prevention efforts should attend to how individuals achieve high levels of daily alcohol consumption through participation in both single drinking occasions and multiple consecutive occasions. This may suggest new intervention approaches and provide insights into the mechanisms driving success or failure of current approaches.

Future research should seek further evidence on who is participating in the occasion types described above, and which occasion- and individual-level characteristics are associated with lower and higher consumption levels within each type. It can also explore how individuals combine occasions across days, weeks and months to reach different levels of alcohol consumption. Using this information, researchers can also examine how to tailor interventions to the drinking occasions or target populations or individuals. Greater attention to mixed-trade drinking occasions as a general phenomenon is needed to understand the associated contexts, participants and practices.

## Conclusion

Alcohol consumption levels vary substantially within and across fifteen occasion types in Great Britain. Heavy drinking is not concentrated in particular occasion types and is instead common across all types. However, moderate drinking is also common in some occasion types of that are often characterised as heavy drinking practices. Mixed-trade drinking occasions are particularly likely to involve heavy consumption and merit greater attention from researchers, policy-makers and practitioners.

## Data Availability

The dataset used in this study is a commercial product owned by Kantar and licensed for use by the research team. Data are therefore not publicly shareable but there is agreement that data, analyses and results can be checked by competent independent reviewers as part of a confidential review process.

