## Appendix A1A for "Alcohol consumption and heavy episodic drinking within different types of drinking occasion in Great Britain: An event-level latent class analysis"

### 1. Weighting procedure

The use of quota sampling may lead to selective samples given that the probability of selecting individuals is unknown. The researcher can employ post-stratification weighting to adjust for differences between a targeted population and the observed sample characteristics. Weighting consists of dividing the sample population into post-stratification groups defined by specific control variables (age, sex, region, etc.) and applying a multiplicative factor so that the distribution of control variables for each subgroup resembles that of the target population. In practice, higher weights are computed and assigned to groups of individuals that are under-represented in the sample and smaller weights are assigned to over-represented groups. There are two common methods for computing sample weights.[1] When the joint probabilities of the target variables are known, *cell-level* weights are calculated for each interlocking cell (e.g. the proportion of women over 65 years old, living in Scotland, and with social grade C1) to achieve the corresponding targets. If the joint probabilities are unknown, an iterative process known as *raking* or *rim weighting* is employed to force the marginal distributions of auxiliary variables (strata) to conform to the joint distribution of the targeted population. Given the non-probabilistic sample design of Kantar Alcovision and the over-representation of targeted categories such as Scotland and 18-24 year old individuals, a raking approach is used in the present analysis.

The general procedure of raking is the following: a weight is applied to each individual in the sample such that the weighted distribution of the first control variable matches the distribution of the same variable in the specified target population. Subsequently, an algorithm readjusts the (weighted) distribution of the second variable to match the target population. This is then repeated for all of the other variables considered. Finally, the adjustment process is reiterated N times until the marginal distribution of all control variables has been perfectly matched with the targets. An advantage of using this raking approach is to reduce bias (i.e. deviation between sample and population means of observed characteristics). However, this may come with a penalty as weight calibration may also lead to an increase in the standard error of sample means.[1] Nevertheless, the benefits of reducing the bias is generally believed to outweigh the cost of an increase in sampling error.[2]

The present analysis uses raking to match the UK Census population profile on three dimensions: social grade, geographic region, and age-sex groups. To avoid weights with very high values, we follow,[3] who suggest collapsing categories of the control variables such that each category adds up to at least 5% of the population units. The raking procedure is conducted in Stata (version 15) with the command *ipfraking* implemented by Kolenikov.[1]

### 2. Latent class model fitting results

*Figure A1.1: Latent class model fit statistics for off-trade only models with two to eight classes.*

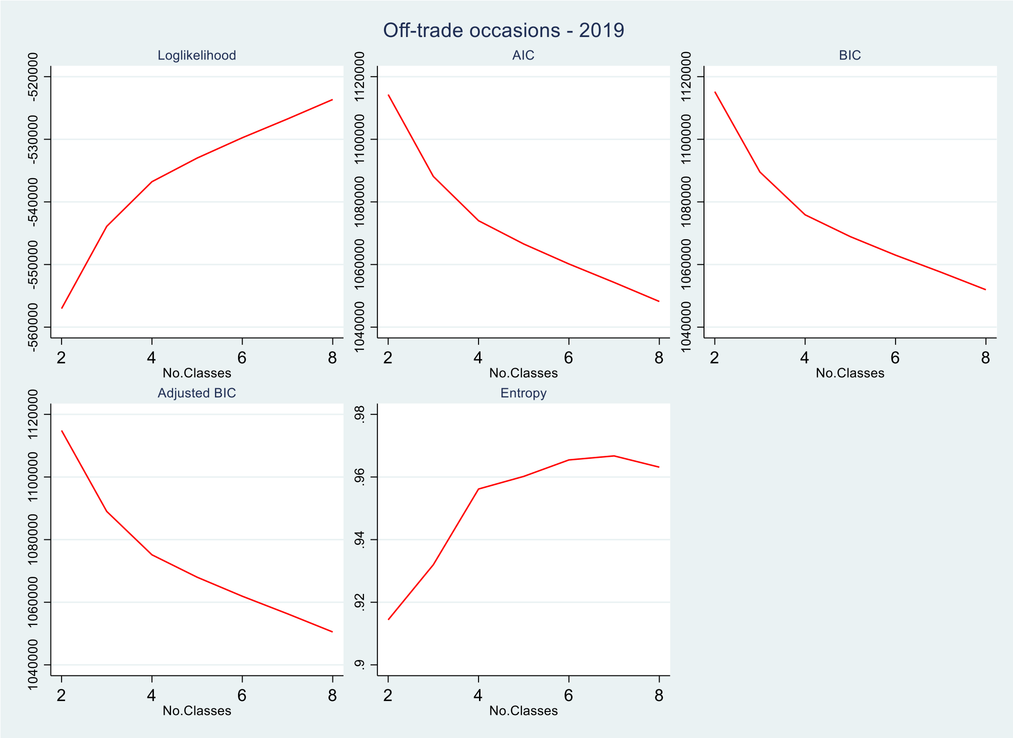


*Figure A1.2: Latent class model fit statistics for on-trade only models with two to eight classes.*

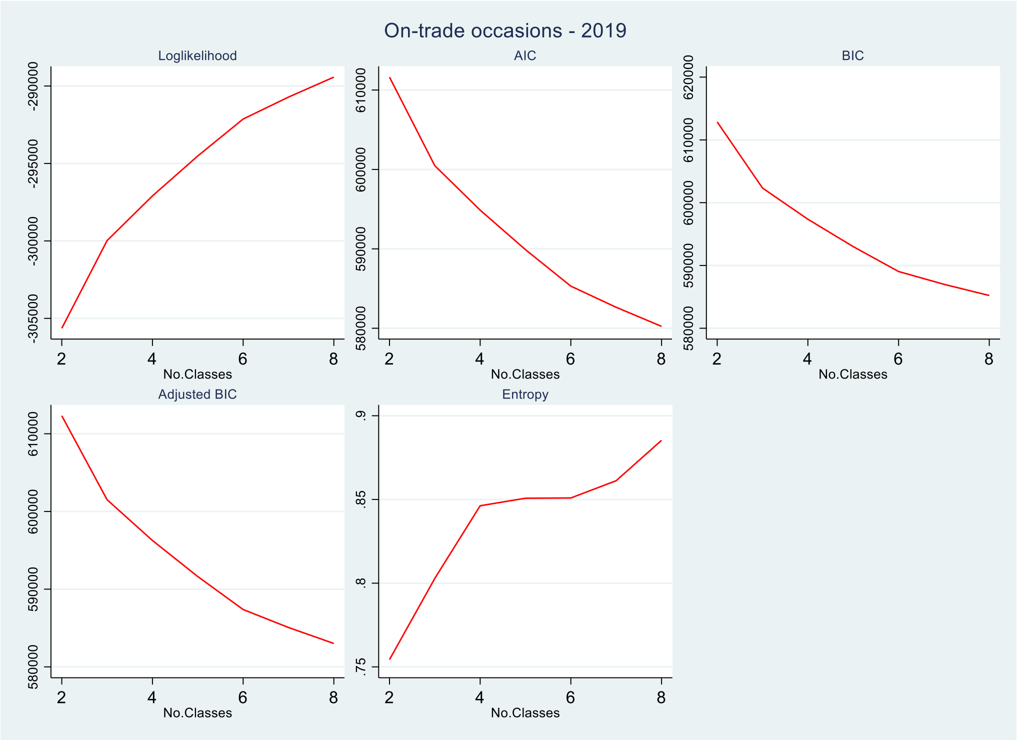

*Figure A1.3: Latent class model fit statistics for mixed-trade models with two to eight classes.*

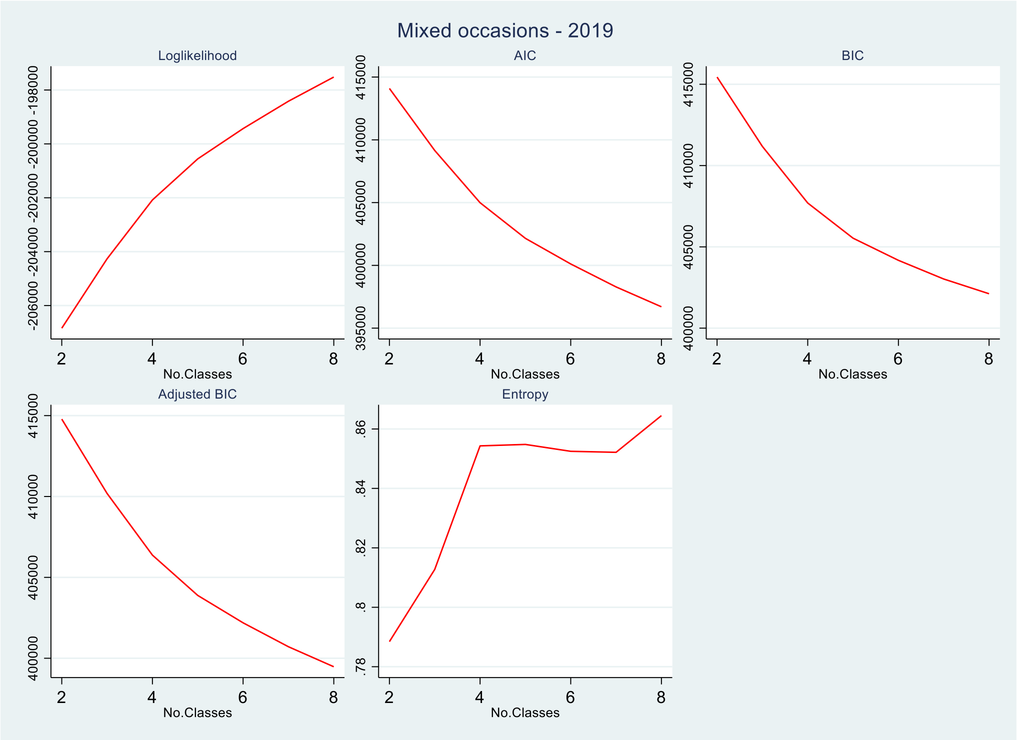

*Table A1.1 - statistical tests of model restrictions for k vs k-1 classes*

| Trade sector | Number of Classes | Number of free parameters | Loglikelihood | AIC | BIC | Adjusted-BIC | Entropy | Test type (p-value) Vuong-Lo-Mendell-Rubin | Lo-Mendell-Rubin adjusted LRT[1] | Bootstrapped LRT[1] |
|---|---|---|---|---|---|---|---|---|---|---|
| Off-trade only | 2 | 113 | -557031 | 1114289 | 1115222 | 1114863 | 0.914 | 0.333 | 0.333 | <0.001 |
| Off-trade only | 3 | 170 | -543902 | 1088145 | 1089549 | 1089009 | 0.932 | <0.001 | <0.001 | <0.001 |
| Off-trade only | 4 | 227 | -536775 | 1074003 | 1075878 | 1075156 | 0.956 | <0.001 | <0.001 | <0.001 |
| Off-trade only | 5 | 284 | -532998 | 1066565 | 1068910 | 1068008 | 0.960 | <0.001 | <0.001 | <0.001 |
| Off-trade only | 6 | 341 | -529753 | 1060189 | 1063005 | 1061921 | 0.965 | <0.001 | <0.001 | <0.001 |
| Off-trade only | 7 | 398 | -526748 | 1054291 | 1057578 | 1056313 | 0.967 | <0.001 | <0.001 | <0.001 |
| Off-trade only | 8 | 455 | -523643 | 1048196 | 1051954 | 1050508 | 0.963 | <0.001 | <0.001 | <0.001 |
| On-trade only | 2 | 173 | -305640 | 611625 | 612866 | 612316 | 0.754 | <0.001 | <0.001 | <0.001 |
| On-trade only | 3 | 260 | -299976 | 600472 | 602336 | 601510 | 0.803 | <0.001 | <0.001 | <0.001 |
| On-trade only | 4 | 347 | -297094 | 594882 | 597370 | 596267 | 0.846 | <0.001 | <0.001 | <0.001 |
| On-trade only | 5 | 434 | -294514 | 589896 | 593008 | 591629 | 0.851 | <0.001 | <0.001 | <0.001 |
| On-trade only | 6 | 521 | -292132 | 585305 | 589041 | 587385 | 0.851 | <0.001 | <0.001 | <0.001 |
| On-trade only | 7 | 608 | -290714 | 582644 | 587003 | 585071 | 0.861 | <0.001 | <0.001 | <0.001 |
| On-trade only | 8 | 695 | -289426 | 580241 | 585224 | 583015 | 0.885 | <0.001 | <0.001 | <0.001 |
| Mixed-trade | 2 | 207 | -206840 | 414094 | 415443 | 414785 | 0.788 | <0.001 | <0.001 | <0.001 |
| Mixed-trade | 3 | 311 | -204265 | 409152 | 411179 | 410191 | 0.813 | <0.001 | <0.001 | <0.001 |
| Mixed-trade | 4 | 415 | -202081 | 404993 | 407697 | 406379 | 0.854 | <0.001 | <0.001 | <0.001 |
| Mixed-trade | 5 | 519 | -200558 | 402153 | 405536 | 403887 | 0.855 | <0.001 | <0.001 | <0.001 |
| Mixed-trade | 6 | 623 | -199433 | 400113 | 404173 | 402193 | 0.852 | 0.533 | 0.534 | <0.001 |
| Mixed-trade | 7 | 727 | -198416 | 398287 | 403025 | 400715 | 0.852 | 0.768 | 0.768 | <0.001 |
| Mixed-trade | 8 | 831 | -197520 | 396703 | 402119 | 399478 | 0.864 | 0.819 | 0.819 | <0.001 |

[1]Loglikelihood ratio test

*Table A1.2: Analysis of misclassification error*

| | | Probability that occasion belongs to assigned class | | | |
|---|---|---|---|---|---|
| **Occasion type** | **N** | **Mean** | **SD** | **Min** | **Max** |
| Quiet drink at home alone | 7,700 | 0.99 | 0.06 | 0.44 | 1.00 |
| Family time at home | 3,946 | 0.94 | 0.11 | 0.45 | 1.00 |
| Evening at home with partner | 8,498 | 0.99 | 0.04 | 0.45 | 1.00 |
| Off-trade get together | 8,351 | 0.96 | 0.10 | 0.50 | 1.00 |
| Meeting friends at the pub | 1,679 | 0.86 | 0.15 | 0.36 | 1.00 |
| Male friends at the pub | 1,080 | 0.93 | 0.13 | 0.31 | 1.00 |
| Quiet drink at the pub | 1,310 | 0.92 | 0.13 | 0.39 | 1.00 |
| Big night out | 824 | 0.91 | 0.15 | 0.28 | 1.00 |
| Extended occasion (on-trade) | 1,658 | 0.89 | 0.16 | 0.27 | 1.00 |
| Family meal | 1,013 | 0.90 | 0.14 | 0.37 | 1.00 |
| Meal with friends | 966 | 0.85 | 0.17 | 0.35 | 1.00 |
| Going out with partner | 1,065 | 0.98 | 0.07 | 0.36 | 1.00 |
| Big night out with pre-drinking | 1290 | 0.91 | 0.13 | 0.40 | 1.00 |
| Quiet drink at home and with friends… | 1735 | 0.93 | 0.13 | 0.36 | 1.00 |
| Extended occasion (mixed trade) | 1974 | 0.91 | 0.14 | 0.38 | 1.00 |

*Table A1.3: Levels and distribution of alcohol consumption and heavy drinking occasions across occasion types (latent classes) for women.*

| | | % of occasions | | Units consumed in occasion | | | | | | | | | % of total consumption | | | Heavy drinking occasions (HDOs) | |
|---|---|---|---|---|---|---|---|---|---|---|---|---|---|---|---|---|---|
| | | | | Total | | | Off-trade | | | On-trade | | | | | | | |
| **Trade sector** | **Occasion type** | **All** | **In trade sector** | **Mean** | **SD** | **Median** | **Mean** | **SD** | **Median** | **Mean** | **SD** | **Median** | **Total** | **Off-trade** | **On-trade** | **% of type[1]** | **% of all HDOs** |
| Off-trade only | Quiet drink at home alone | 17.8 | 24.2 | 12.5 | 7.1 | 9.8 | 12.5 | 7.1 | 9.8 | - | - | - | 7.5 | 10.6 | - | 25.4 | 12.5 |
| | Family time at home | 9.4 | 12.8 | 14.0 | 7.6 | 10.8 | 14.0 | 7.6 | 10.8 | - | - | - | 6.7 | 9.5 | - | 32.3 | 8.4 |
| | Evening at home with partner | 28.6 | 39 | 11.6 | 6.4 | 9.7 | 11.6 | 6.4 | 9.7 | - | - | - | 11.3 | 16.1 | - | 25.6 | 20.4 |
| | Off-trade get together | 17.5 | 23.9 | 16.5 | 10.0 | 12.9 | 16.5 | 10.0 | 12.9 | - | - | - | 31.0 | 44.0 | - | 49.0 | 23.9 |
| On-trade only | Meeting friends at the pub | 3.1 | 17.8 | 11.0 | 5.7 | 9.5 | - | - | - | 11.0 | 5.7 | 9.5 | 2.2 | - | 7.4 | 32.2 | 2.8 |
| | Male friends at the pub | 0.1 | 0.7 | 12.7 | 3.2 | 12.3 | - | - | - | 12.7 | 3.2 | 12.3 | 0.1 | - | 0.5 | 57.1 | 0.2 |
| | Quiet drink at the pub | 0.9 | 5.1 | 9.9 | 3.2 | 9.5 | - | - | - | 9.9 | 3.2 | 9.5 | 0.3 | - | 1.1 | 16.3 | 0.4 |
| | Big night out | 1.6 | 8.9 | 14.2 | 7.3 | 12.0 | - | - | - | 14.2 | 7.3 | 12.0 | 3.7 | - | 12.4 | 58.8 | 2.6 |
| | Extended occasion (on-trade) | 2.3 | 13.0 | 20.3 | 10.9 | 17.1 | - | - | - | 20.3 | 10.9 | 17.1 | 8.8 | - | 29.6 | 73.7 | 4.7 |
| | Family meal | 3.1 | 17.6 | 10.5 | 5.1 | 9.5 | - | - | - | 10.5 | 5.1 | 9.5 | 1.0 | - | 3.4 | 17.6 | 1.5 |
| | Meal with friends | 3.4 | 19.4 | 11.0 | 5.7 | 9.7 | - | - | - | 11.0 | 5.7 | 9.7 | 1.7 | - | 5.6 | 25.4 | 2.4 |
| | Going out with partner | 3.1 | 17.6 | 10.9 | 5.1 | 9.4 | - | - | - | 10.9 | 5.1 | 9.4 | 1.1 | - | 3.7 | 21.9 | 1.9 |
| Mixed-trade | Big night out with pre-drinking | 2.4 | 26.1 | 18.3 | 9.6 | 15.3 | 9.3 | 7.0 | 7.5 | 9.0 | 6.7 | 7.5 | 8.8 | 6.6 | 14.3 | 82.3 | 5.5 |
| | Quiet drink at home and with friends… | 2.6 | 27.7 | 13.7 | 7.5 | 11.1 | 7.6 | 6.3 | 5.7 | 6.1 | 4.5 | 5.0 | 3.5 | 2.8 | 5.3 | 62.7 | 4.5 |
| | Extended occasion (mixed trade) | 4.3 | 46.2 | 19.7 | 11.1 | 15.9 | 11.6 | 9.5 | 8.2 | 8.1 | 6.7 | 6.0 | 12.3 | 10.4 | 16.8 | 69.8 | 8.3 |

*Table A1.4: Levels and distribution of alcohol consumption and heavy drinking occasions across occasion types (latent classes) for men.*

| | | % of occasions | | Units consumed in occasion | | | | | | | | | % of total consumption | | | Heavy drinking occasions (HDOs) | |
|---|---|---|---|---|---|---|---|---|---|---|---|---|---|---|---|---|---|
| | | | | Total | | | Off-trade | | | On-trade | | | | | | | |
| Trade sector | Occasion type | All | In trade sector | Mean | SD | Median | Mean | SD | Median | Mean | SD | Median | Total | Off-trade | On-trade | % of type[1] | % of all HDOs |
| Off-trade only | Quiet drink at home alone | 21.1 | 32.2 | 15.7 | 7.8 | 13.2 | 15.7 | 7.8 | 13.2 | - | - | - | 14.0 | 21.2 | - | 28.6 | 15.2 |
| | Family time at home | 9.2 | 14.1 | 15.0 | 7.5 | 12.3 | 15.0 | 7.5 | 12.3 | - | - | - | 6.4 | 9.7 | - | 32.3 | 7.5 |
| | Evening at home with partner | 19.4 | 29.7 | 14.6 | 6.6 | 12.0 | 14.6 | 6.6 | 12.0 | - | - | - | 11.5 | 17.3 | - | 29.4 | 14.4 |
| | Off-trade get together | 15.7 | 24.0 | 19.1 | 9.7 | 16.0 | 19.1 | 9.7 | 16.0 | - | - | - | 21.8 | 32.9 | - | 47.3 | 18.6 |
| On-trade only | Meeting friends at the pub | 4.3 | 18.2 | 13.6 | 6.0 | 11.4 | - | - | - | 13.6 | 6.0 | 11.4 | 3.6 | - | 10.6 | 45.2 | 4.8 |
| | Male friends at the pub | 4.7 | 20.2 | 14.6 | 6.0 | 12.5 | - | - | - | 14.6 | 6.0 | 12.5 | 4.9 | - | 14.4 | 55.7 | 6.6 |
| | Quiet drink at the pub | 4.4 | 19.0 | 12.4 | 4.6 | 11.2 | - | - | - | 12.4 | 4.6 | 11.2 | 1.9 | - | 5.6 | 24.7 | 2.8 |
| | Big night out | 1.0 | 4.4 | 18.9 | 9.4 | 17.2 | - | - | - | 18.9 | 9.4 | 17.2 | 1.7 | - | 5.1 | 57.7 | 1.5 |
| | Extended occasion (on-trade) | 3.6 | 15.2 | 22.0 | 10.8 | 19.2 | - | - | - | 22.0 | 10.8 | 19.2 | 8.1 | - | 23.9 | 66.4 | 5.9 |
| | Family meal | 1.7 | 7.5 | 13.4 | 6.3 | 11.1 | - | - | - | 13.4 | 6.3 | 11.1 | 0.5 | - | 1.6 | 16.1 | 0.7 |
| | Meal with friends | 1.4 | 5.8 | 15.3 | 7.6 | 12.2 | - | - | - | 15.3 | 7.6 | 12.2 | 0.6 | - | 1.9 | 21.5 | 0.7 |
| | Going out with partner | 2.3 | 9.9 | 13.4 | 5.9 | 11.4 | - | - | - | 13.4 | 5.9 | 11.4 | 1.0 | - | 3.0 | 23.1 | 1.3 |
| Mixed-trade | Big night out with pre-drinking | 2.3 | 20.4 | 21.9 | 9.8 | 19.6 | 10.5 | 8.1 | 8.0 | 11.3 | 7.3 | 10.0 | 6.5 | 4.9 | 9.7 | 84.6 | 4.9 |
| | Quiet drink at home and with friends… | 5.2 | 46.8 | 16.8 | 8.1 | 14.6 | 8.2 | 6.6 | 6.4 | 8.6 | 5.5 | 7.4 | 8.2 | 6.1 | 12.2 | 66.2 | 8.7 |
| | Extended occasion (mixed trade) | 3.7 | 32.9 | 23.3 | 10.7 | 21.2 | 13.1 | 9.4 | 10.7 | 10.2 | 7.7 | 8.5 | 9.4 | 8.0 | 12.2 | 69.4 | 6.4 |
